# A mapping review of good practices of participatory research for an impactful collaboration in disabilities studies

**DOI:** 10.1101/2024.09.18.24313890

**Authors:** Maëlle Corcuff, Rania Jribi, Guillaume Rodrigue, Marie-Eve Lamontagne, Émilie Raymond, Philippe S. Archambault, François Routhier

## Abstract

**Introduction:** Participatory research is particularly relevant to understanding the challenges faced by people with disabilities (PWDs), as it actively involves them as partners, enabling methodologies to be better adapted to lived realities and producing more relevant and applicable results. By reducing systemic barriers and promoting inclusion, this approach improves understanding and consideration of the specific needs of PWDs in research. Yet, studies have identified hurdles associated with this approach, prompting questions about how organizations portray PWDs, the dynamics among research stakeholders, the distribution of decision-making power, and the actual impact of research on its partners.

**Aim:** This study aims to identify the factors that influence the process and results of participatory research in the field of disability studies

**Methods:** We conducted a mapping review following the PRISMA-ScR guidelines, and analysis the results according to the input-throughput-outcomes Bergen model

**Results:** This study identifies partners skills and training, power sharing and benefits of active involvement as facilitators of participatory research. On the other hand, contextual challenges, and lack of guidance are reported as obstacles.

**Conclusion:** This study provides insight into how the various facilitators and obstacles to participatory research and its different processes interact to produce positive, valid and rigorous results.

## Introduction

People living with disabilities (PWDs) are confronted daily with multiple systemic barriers in their physical and social environment, threatening their full participation in society, such as access to health services, employment, leisure activities or education [1–5]. Yet the establishment of an inclusive society is at the heart of political and economic concerns, to enable all individuals to exercise their rights and participate fully. The research community is no exception in building a more inclusive society. Over the last few decades, this topic has become of interest to researchers, with the aim of developing innovative solutions implemented by and for a diversity of stakeholders, such as disabled people themselves and the organizations representing them [6].

Research in the field of disabilities studies has undergone a significant evolution in recent years, with more use of participatory research approaches. Participatory research aims to develop knowledge to improve and solve issues experienced by PWDs, with them as active partners rather than as research subjects [7]. This type of research involves close collaboration between PWDs, organizations representing these people and academic researchers in the process [8]. This can include different methodological approaches and terminologies such as co-production, integrated knowledge production, community-based participatory research, partnership research or action research [9].

Participatory research with PWDs, organizations that represent them, advocacy groups or services providers, has expanded considerably in recent years. Numerous studies have demonstrated the beneficial influence of this type of research-on-research processes [9, 10] as well as on the adoption of results through changes in practices [10]. Participatory research offers several advantages, such as greater social relevance of research, interventions, practices and instruments that are better adapted to needs, and greater potential for appropriation of results by knowledge users [9]. In the field of disabilities, participatory research is also seen as a way for PWDs and organizations to play an active role in identifying research priorities and in the processes themselves [11]. Indeed, they can actively participate in every stage of a research project, from the identification of the problem to the mobilization of the knowledge acquired as part of the study [12].

However, studies have highlighted obstacles associated with participatory research. Indeed, participatory approaches can represent significant challenges for researchers or partners (both organizations and individuals) in terms of the time, energy, and skills they require [6]. This raises several questions about the way in which organizations represent PWDs, the nature of relationships between research players, the way in which decision-making is shared and how research has a real impact on partners [13].

In order to shed light on the obstacles and facilitators of participatory research, we have sought to circumscribe the knowledge related to the different aspects and practices of participatory research in disability studies. Thus, this mapping review aims to identify the factors that influence the process and results of participatory research in the field of disability studies.

## Methods

We conducted a mapping review, based on the sequential stages of the systematic mapping process [14]. The findings were reported according to the scoping review extension of the preferred reporting items for systematic review and Meta-Analyses (PRISMA-ScR) guidelines [14]. A mapping review aims to describe and categorize knowledge within a topic known scope to identify gaps in literature [15]. Thus, the main reason of conducting this mapping review was to describe and categorize knowledge about participatory research in the field of disability studies [14, 15].

### Literature search

A search was conducted in twelve electronic databases using a search strategy to ensure that all articles dealing with the topic were identified (ABI Inform, Ageline, Anthropology Plus, CINAHL Plus with Full Text, International Bibliography of the Social Sciences (IBSS), Medline, PsycInfo, Sociological Abstracts, Social Science Full Text, Social Service Abstracts, Women’s Studies International, Worldwide Political Science Abstracts). Specific keywords and MESH terms were used, such as collaborat* or partner* or participatory or community based or community centered AND guidelines as topic or practice guidelines as topic or meta-analysis as topic or meta-analysis or models, theoretical or models, organizational. The different research scripts are presented in Appendix 1. We limited our search to articles published in English or in French between 2008 and 2023.

### Study selection

The review method was based on two steps: review of the title and abstract and a full text review by two independent reviewers (MC and GR). Conflicts were discussed and resolved with a third author (FR). The following inclusion criteria were established to select the articles: (1) Directly related to disabilities (2) Written in English or French, (3) Presents research results (must be about the results of a scientific process), (4) Presents elements of a PR process (e.g., model, good practices, or effects of PR) and (5) Since 2008.

### Data extraction and analysis

Since mapping review has a broad focus, but with limited data extracted from the included papers, we only extracted descriptive information about the studies and applied predefined codes [14]. Thus, after identifying relevant articles, one author (GR) extracted data in an Excel grid with items determined by the research team. The items were as follow: (1) Objectives of the article, (2) Research question, (3) Population studied, (4) Typology used and definition of PR, (5) Steps of the project, (6) Actors involved and their role, (7) Facilitators of the process, (8) Obstacles to the process, (9) Good practices identified and (10) Issues encountered.

To classify the extracted results, we used a deductive approach following the Bergen model [16], as prescribed by the methodology for mapping reviews [14].The Bergen Model is a theorical framework developed and tested in health promotion studies. It presents three important categories that interact in a cyclical process within the context of collaboration [16–19]. The **input** category includes partners resources (personal factors related to researchers, partners’ readiness, benefits, and technical support to partners), financial resources such a financial support that encourage recruitment of additional inputs and the mission (objectives, setting a common ground for the research)[18]. In the **throughput** category, multiples elements of inputs enter the collaboration process. They interact positively or negatively depending on leadership, communication, roles and structure [18]. The **output** category of the collaboration are additive results (additive results within the project and within partners), synery (facilitators or obstacles of great achievement as a team), antagonistic results (in this study what could be an obstacle for an additive result is a facilitator for an antagonistic results)[18].

## Results

### Characteristics of included studies

42 studies were included in the review. Figure 1 illustrates the PRISMA selection process for the articles. Seventeen of the selected studies focused on developmental disorders including studies on intellectual disabilities (n=10), autism spectrum disorder (n=5) and other developmental disorders (n=2). Other studies covered disabilities related to mental disorders (n=9), dementia (n=4) and disabilities in general (n=4). Two (n=2) studies considered multiple types of disabilities and others covered physical disabilities, invisible disabilities (e.g., epilepsy), cognitive impairments, pain, rheumatism, and other health problems. Characteristics of studies are reported in appendix 2. We have used Bergen’s model to examine a range of participatory research methodologies across various studies. Identified facilitators and barriers were classified in three categories as participatory inputs, throughput and outcomes.

**Figure 1.**
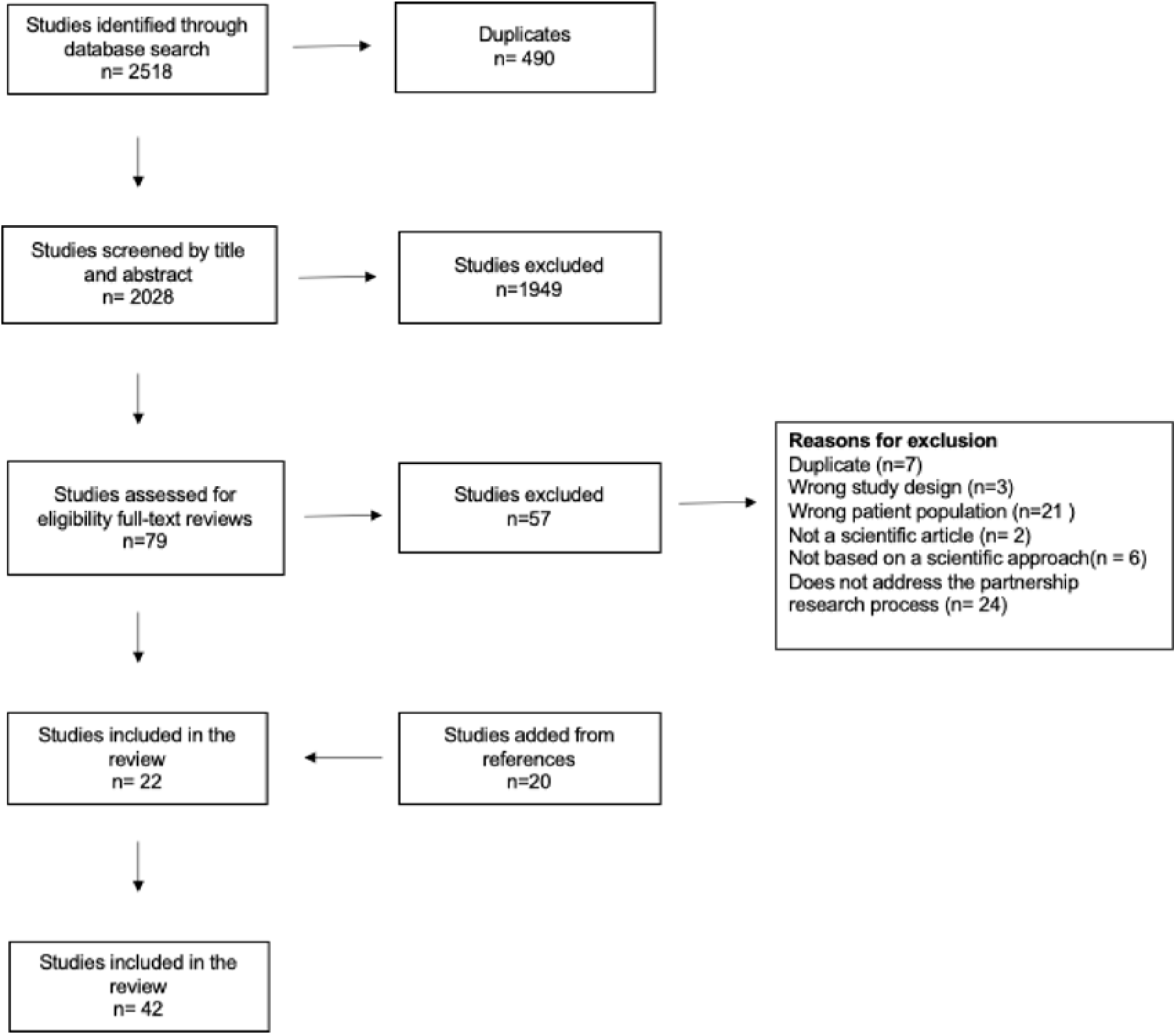
PRISMA flowchart

#### 1. Facilitators

The Bergen’s model is based on inputs, throughputs, and outputs [20]. In this review, we extracted various elements from participatory research studies and used Bergen’s model to classify these elements accordingly. We categorized the factors entering collaboration as inputs, such as partners’ skills, training, research purposes, expectations, and funding. We examined the activities occurring during the collaboration process and classified them as throughputs, including power sharing, researcher collaboration, relationship building, skills development, and engagement support strategies. Finally, we identified the outcomes resulting from the collaborative process as outputs, which include benefits, active involvement, and impacts on methods and practices. This section presents these factors and elements, highlighting how they facilitate successful collaborative functioning and enhance the likelihood of achieving favorable outcomes. Figure 2 illustrates the various facilitators of participatory research that emerged from the studies included in this review.

**Figure 2.**
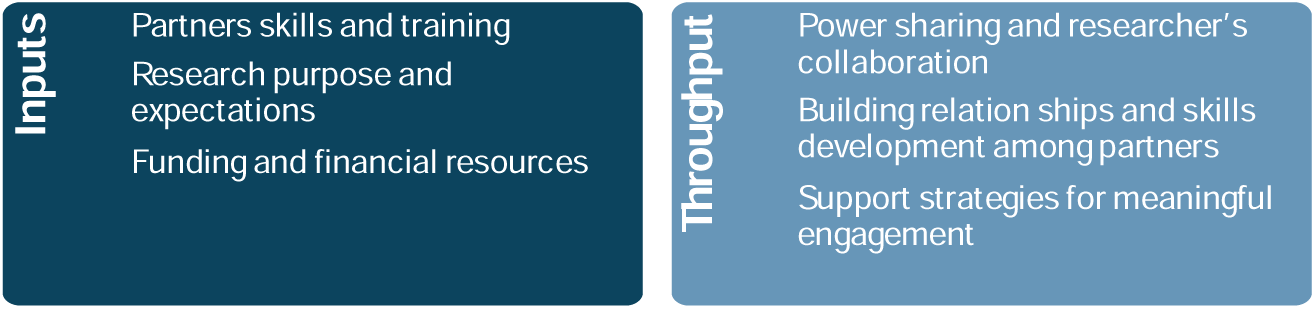
Facilitators of participatory research according to Bergen Model

##### 1.1. Inputs

###### Partners skills and training

This category refers to the diverse expertise and abilities that various partners bring to collaboration. It includes subcategories such as research training, skills development for participants, recruitment of multiple partners, and the inclusion of PWDs. In disability studies, various stakeholders – including researchers and PWDs – contribute diverse resources, such as skills, expertise, and personal characteristics that must interfere in interactions or contributions[16]⍰. For instance, previous studies suggest that researchers who have previous experience in participatory research tend to collaborate more [13, 21, 22]⍰. Age-related factors also play a significant role. Desai et al. (2019) found that younger researchers often find it easier to communicate with their partners[23]⍰. Furthermore, studies indicate that when researchers invest time in understanding their partners from different communities, align their visions, and adopt a collaborative research approach, it positively impacts participatory research and encourages active participant contribution[24, 25]⍰. Researchers lacking experience in participatory research benefit significantly from additional support in navigating participatory methods and understanding the foundational literature of this research approach[26]⍰. Therefore, introducing young researchers to the principles and methodologies of participatory research emerges as a promising solution[26]⍰.

**Training for researchers.** Abma et al. (2009) also advocate that researchers should undergo training in participatory research management through courses that facilitate mutual learning between researchers and partners [27]. De Wit et al. (2015) similarly emphasized the need for additional guidance for researchers in conducting participatory research [26]. To enhance their skills, researchers could participate in courses specifically focused on participatory research methods and empowerment strategies. Partners also play a crucial role by offering insights into community needs and effective engagement methods [27, 28]. Abma et al. (2009) proposed the development of courses tailored to both partners and professionals, promoting reciprocal learning and collaboration [27]. Such training initiatives are described as essential in bridging the gap between research and practical application [22]. Additionally, Desai et al. (2019) highlighted the potential of informal gatherings where community partners and researchers interact, share experiences, and build relationships, which can be more impactful than traditional academic conferences for fostering effective participatory research [23].

**Skills development and training for participants.** To enhance participatory research, Conder et al. (2011) advocated identifying individuals with relevant skills, emphasizing their effective contribution to group discussions [29]. Roberge-Dao et al. (2019) highlighted the ease with which partners with clinical experience integrate into research projects. However, this approach may overlook skills development for those lacking these abilities [29, 30]. Integrating training programs in participatory research is suggested to foster skills development [13], ensuring diverse perspectives and meaningful engagement in projects [13, 25, 27, 28, 31]. De Wit et al. (2015) argued that training partners demonstrates respect, enhancing their familiarity with research processes and decision-making [26]. Mutual learning and exchanging experiences can motivate partners and strengthen their team cohesion [26].

Investing in partner training is critical, especially for community members new to research [8, 29, 32]. PWDs in pivotal roles may require extensive training [8]. According to Ottmann et al. (2009), ongoing capacity building is essential for partners, even in project culmination stages. Camden et al. (2015) used research assistants as mentors to enhance partners’ research skills. De Wit et al. (2015) highlighted the importance of fostering personal interactions among partners during training to promote mutual support. Studies have stressed the need to tailor training to partners’ specific needs. For team members managing chronic pain and fatigue, training should include frequent breaks and sessions under 90 minutes. Thoft et al. (2020) Balanced Participation Model emphasized continuous skill training for individuals affected by dementia, advocating immediate application of acquired skills in ongoing projects. Tanner’s model addressed training retention challenges in dementia care, suggesting simplified documents and memory aids for post-training recall [33]. Bigby and Frawley (2010) recommended focusing on partners’ strengths and addressing research challenges within the research context rather than formal training for individuals with intellectual challenges [34]. Healthcare providers may benefit from immersion in active research settings for developing crucial research-related skills [21].

**Partners recruitment.** Involving multiple partners was often viewed advantageous [35]. Collaborating with various partners helped balance the participatory research, encouraging researchers to be more attentive to their needs while preventing them from being overwhelmed by responsibilities [27]. Recruiting additional participants, especially for groups with progressive disabilities like dementia, becomes crucial during participative research projects to manage departures that may occur due to the inability to continue the research process [25]. Diversifying the array of partners can help mitigate the impact of departures [25]. Some researchers highlighted the benefits of having a broad network of disability organizations and a diverse community of PWDs, as it was seen as providing a wide pool of talent and expertise for participatory research [36]. Ehde et al. (2013) observed that collaborating with well-structured organizations experienced in participatory research facilitates the process [11].

**Inclusion of PWDs.** Empowered PWDs contributed effectively as research partners, providing valuable insights [30]. Ensuring meaningful involvement of PWDs in all research stages was viewed as enriching but required accommodations for mobility, pain, fatigue, and other needs [8, 31]. Budgeting for wheelchair-accessible transportation and specialized training materials is essential [8]. To include individuals preferring non-verbal communication, diverse participation (ex. Planning an action worksheet, sticker voting) and organizational (ex. Money list, place to store things) strategies are deemed as crucial [37]. Despite concerns, individuals with intellectual disabilities can contribute actively when their viewpoints are acknowledged in collaborative processes [34, 38]. In addition, empowering them with information about their capabilities was described as fostering informed decision-making [38]. Collaborating with individuals affected by dementia necessitates ongoing adjustments to accommodate changing abilities [25], emphasizing the need to adapt methodologies throughout the study.

*<u>Research purpose and expectations</u>*. This outlines the reason for collaboration, detailing the objectives and anticipated outcomes of the project. Studies emphasized the critical role of research questions in shaping participatory research. Many studies indicated that agreement on the research question promoted a smoother process [13, 32, 37, 39–41]. Collaborative identification of these questions enhanced partners’ ownership of the project [13, 22, 40]. Establishing a shared project objective and unified vision helped prevent conflicts and fosters transparency throughout the research stages [41]. This alignment nurtured a cohesive research environment [37]. Early establishment of clear and realistic expectations has been recommended to facilitate achieving research outcomes and meeting partners’ needs [13, 22]. Transparency before starting the research process is seen as crucial, acknowledging that participatory research may have limited impact on a large scale [42]. Setting boundaries and timelines has allowed partners to align efforts toward common objectives [22, 30, 43], providing a framework for managing the project [30]. Visual exercises at project inception can clarify and reinforce these shared objectives [30].

*<u>Funding and financial resources</u>*. Funding bodies have played a pivotal role in shaping research directions and methodologies to advance participatory research. Fletcher-Watson et al. (2019) suggested that funding bodies should mandate community consultation in all proposals, with evaluators assessing proposed participatory activities [43]. Stolee et al. (2011) proposed refining funding proposal evaluation criteria to promote the engagement of healthcare providers in collaborative research [44]. Moreover, Camden et al. (2015) recommended restructuring funding models to encourage sustained involvement across multiple projects rather than focusing solely on single-project funding [13].

##### 1.2. Throughput

*<u>Power sharing and researchers’ collaboration</u>*. This highlights the importance of distributing responsibilities among all involved partners to ensure equitable participation and effective teamwork on the project. Participatory research involves multiple partners interacting throughout the process. Several studies focused on participatory research [24, 45, 46], highlighted power-sharing as a significant challenge [46]. Facilitators to collaboration included structured participatory frameworks such as regular meetings to align strategies toward shared goals, assessing beliefs about disability, ensuring information accessibility, and evaluating partner engagement and influence in meetings [45]. Morton (2012) emphasized the value of peer debriefing sessions to improve work accuracy, while consultation among researchers guides participatory research actions [24].

*<u>Building relationships and skills development among partners</u>*. This category emphasizes the importance of collaborative connections between partners and their role in enhancing skills and sustaining the partnership. Other studies have questioned how partner relationships influence participatory research progress [26, 27, 30, 31, 36, 37, 45, 46]. Skill development among partners is described as crucial to the participatory process [26, 30, 45]. Regular exchange of experiences and mutual learning are also essential for sustaining participatory research [26]. Participatory research allowed individuals with shared life experiences to connect, fostering new friendships, colleagueship, and mentorships [31, 46]. Many studies indicated that this mutual support enhanced team cohesion and organizational capacity for meetings [26, 30, 31, 36, 37, 39, 45, 46]

*<u>Support strategies for meaningful engagement</u>*. This involves creating and implementing methods and accommodations to facilitate effective involvement from all partners.

Researchers sharing control positively influenced partner engagement [13]. Extensive researcher support is critical for meaningful involvement of PWDs in research teams [13, 27, 29, 30, 36, 37, 44, 47]. This support included careful preparation, sensitivity to individual needs, and consistent communication [47]. Adequate support, including transportation, was essential, as emphasized by Conder et al. (2011)[29]. Providing disability accommodations and regularly assessing partner needs was also vital [30, 46]. Ensuring full inclusion of physically disabled individuals necessitates reasonable accommodations [8, 31]. Accessible spaces and materials were likely key facilitators of engagement [45, 46]. Accommodations such as interpreters and accessible materials further enhanced successful participation [8, 30, 35, 46].

Stolee et al. (2011) proposed strategies to enhance collaborative research among healthcare providers, emphasizing the importance of funding and dedicated time to promote participation in multidisciplinary projects [44]. They underscored the need to foster a research culture in healthcare sectors, advocating for increased awareness and appreciation of participatory research. In addition, Roberge-Dao et al. (2019) offered complementary solutions to address organizational challenges faced by healthcare providers, recommending strategies such as allocating protected research time, providing financial incentives, and prioritizing quality care over quantity for patients within healthcare organizations[22]. These combined approaches aimed to overcome organizational barriers and encourage healthcare providers’ involvement in collaborative research efforts within healthcare settings.

Researchers often compensated partners with disabilities for their participation, acknowledging their expertise and commitment [26, 27, 43]. This compensation is crucial for showing respect and appreciation for their efforts toward project objectives [26, 27, 43] and significantly enhanced engagement [46, 48]. Transparent communication about funding constraints allowed partners to make informed decisions regarding their involvement in uncompensated research activities [30], empowering them to manage their roles effectively.

##### 1.3. Outcomes

*<u>Benefits and active involvement.</u>* The engagement of various partners in participatory research can lead to significant outcomes within the project [11, 13, 23, 27, 28, 34, 40, 41, 49, 50]. Many studies indicated that the active involvement of partners can contribute to identifying relevant research questions, strengthening the credibility of findings, and facilitating the practical application of research outcomes within the target population [13, 27, 46, 49, 50]. This proactive engagement ensured that the research project holds the potential for impactful and meaningful outcomes to benefit the studied population [50].

Engaging in research projects offered benefits beyond financial compensation. Partners gained access to facilities, attend events, and participated in social activities [26, 27]. Researchers may provide equipment and offer co-authorship credit, enhancing academic and employment opportunities [49].

*Impact of collaboration on research methods, partner involvement and inclusive practices*.

This category highlights how collaboration helps align research approaches with the needs of participants, enhancing accessibility and integration.

Collaborating with partners who have relevant lived experiences enabled researchers to refine research language, reducing cultural barriers and resonating better with potential participants [11, 13, 23, 27, 28, 34, 40, 41, 49, 50]. This also facilitated the adaptation and validation of data collection instruments, ensuring alignment with the specific needs of the study population [34, 50, 51]. Moreover, partners’ guidance made research participation more accessible by providing multiple engagement methods [30]. Community partners helped identify and address participation risks often overlooked by researchers, empowering potential participants with a clearer understanding of the potential risks associated with their involvement in the research process [23].

#### 2. Obstacles

This section outlines the obstacles identified in participatory research studies, categorized using the Bergen model. We classified challenges related to partners, such as issues with authority figures, representative organizations, contextual challenges, partner training, and funding difficulties, as elements within inputs. Obstacles such as lack of guidance, management challenges, and difficulties in achieving effective partner involvement were classified as elements interfering within the collaboration process, or throughputs. Finally, we present additional challenges and other factors emerging from various interactions in participatory research studies as outcomes. Figure 3 illustrates the various obstacles of participatory research that emerged from the studies included in this review.

**Figure 3.**
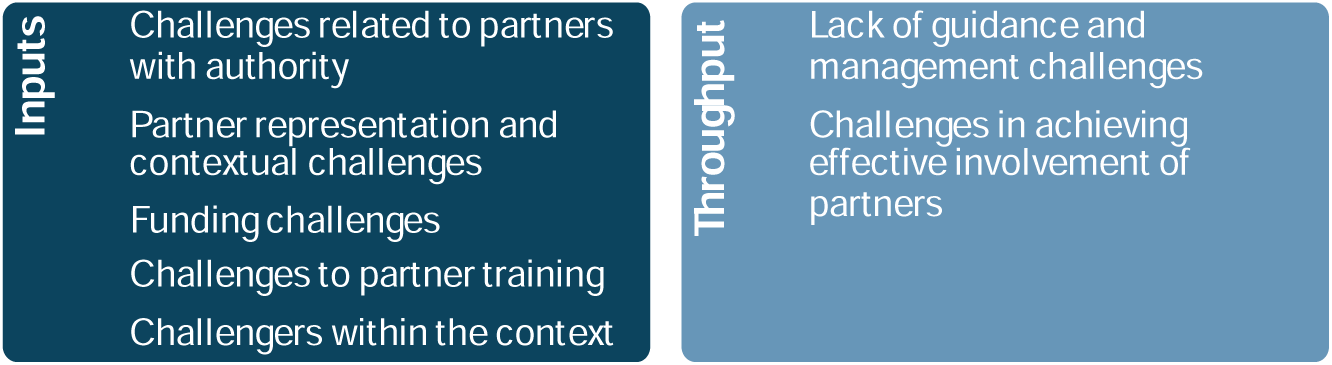
Obstacles of participatory research according to Bergen Model

##### 2.1. Input

###### Challenges related to partners with authority

This category addresses the issue of differing priorities encountered when collaborating with government entities. Minkler et al. (2008) highlighted a challenge when government is involved in participatory research, where the team may face a dilemma between supporting community needs versus those of the government. Bigby and Frawley (2014) mentioned partners with intellectual disabilities expressing concerns about potential implications of discussing government funding for community homes, leading to self-censorship to avoid trouble with the department [34]. One study suggested mistrust and apprehension within the disability community regarding research institutions, posing significant collaboration barriers [23].

Ottmann et al. (2011) shed light on the challenge of identifying specific government branches responsible for pivotal decisions, making it difficult to involve relevant high-level officials in research group meetings. In their work, key decision-makers declined participation, fearing their involvement might compromise their position in broader government processes. In addition, time constraints led senior officials to delegate meeting attendance to subordinates with limited problem-solving abilities [31].

###### Partner representation and contextual challenges

This category refers to the limited number of partners and supportive organizations, as well as the contextual difficulties they face, which impact their involvement and contributions to participatory research. The number of partners played a critical role in balancing power dynamics in research projects [13, 27]. An imbalance between the number of partners and the number of researchers can lead to reduced partner representation in the team [13]. Furthermore, losing partners over time can compromise participatory research, and replacing them mid-study poses significant challenges [51]. Challenges also arose due to limited representation of supportive organizations, constraining the talent pool for participative research involving PWDs [31]. Additionally, gaps in diversity within disability organization networks hindered obtaining comprehensive perspectives and solutions in collaborative research settings [36, 43]. These obstacles strongly impacted the effectiveness and inclusivity of research involving PWDs. Socio-economic factors can influence the ability of community organizations to engage in participatory research. Moule and Davies (2016) highlighted decreased organizational budgets resulting in reduced effectiveness in recruiting study participants. They also faced challenges in obtaining local authority support for recruitment due to lacking necessary documents at the study’s outset [48].

###### Funding challenges

This addresses the difficulties encountered in securing and managing financial resources to fund participatory research project involving PWDs. Community participation can often be viewed by funders as a symbolic gesture, posing a significant challenge for collaborative research [43]. This hurdle was described as impeding the establishment of genuinely collaborative research efforts, which require substantial time and resources. The competitiveness of research proposals may suffer if collaboration isn’t valued by funding bodies [43]. Additionally, funders may hesitate to include PWDs in participatory research, particularly autistic individuals, fearing potential compromise the rigor of the scientific process [43]. This may therefore limit the meaningful involvement of autistic individuals and their families in funded research decisions [43].

Funding timelines also presented challenges for complete partner involvement in research initiatives [13, 26]. Indeed, waiting and delaying project start-up due to approval periods can diminish partners’ motivation and frustrate their engagement, especially when they are involved post-funding, limiting their contribution during crucial project initiation phases [26]. Balancing the needs expressed by funding organizations with those of partners in defining research questions is seen as another dilemma faced by researchers [26, 34, 36]. Bigby and Frawley (2014) illustrated the consequences of excluding partners from defining research questions, resulting in abstract inquiries that may not resonate with the population with intellectual disabilities[34].

Addressing the productivity challenges associated with the research environment in participatory research with PWDs requires inclusive strategies [8]. This may involve integrating training for team members, or even extending the duration of the research project to accommodate tasks, particularly for people with physical or psychiatric symptoms [8, 13]. However, the additional time and resources required for participatory research may reduce its competitiveness in grant applications, particularly if funders do not prioritize community representation within research teams [43]. Uncertainties about research outcomes and timelines can also pose difficulties, exacerbated by budget constraints and final reporting deadlines [29]. Limited budgets may restrict the flexibility to implement partner-suggested protocol changes, while time constraints can hinder adequate preparation of partners for research activities like conducting interviews [29]. Researchers may also face significant workloads to meet project deadlines [27]. Finally, ensuring partner retention until data dissemination was identified as a challenge in short-term funding scenarios unless carefully planned [13, 34].

###### Challenges to partner training

This refers to the obstacles faced in developing the skills of partners involved in the research project. Researchers expressed concerns regarding partner training in participatory research [27, 42]. Abma et al. (2009) highlighted potential negative impacts of partner training, suggesting it may lead partners to adopt a professional status, potentially distancing them from the communities they represent [27]. Additionally, Littlechild et al. (2015) discussed the significant extra costs associated with training partners, adding to challenges within collaborative research projects [42]. These concerns emphasized the need for more nuanced and inclusive approaches to partner training to prevent potential adverse consequences. However, the absence of formal training programs in participatory research could be a significant barrier, particularly in addressing the specific needs of individuals affected by dementia [25]. Disabilities such as attention and memory impairments can limit the capacity of certain individuals, like those with dementia, to fully engage in training programs [25]. Similar challenges were observed among individuals with chronic pain and fatigue, who struggle with extended training sessions lacking adequate breaks [8]. Lack of interest among partners in the research domain can also restrict effective training [34]. These challenges underscored the importance of flexible, adaptive training programs to accommodate diverse needs while balancing the limitations researchers face in resource allocation and project timelines [31].

###### Challenges within the context

This category highlights the contextual difficulties encountered by healthcare providers when participating in research projects. The participatory research context presented numerous challenges, including bureaucratic obstacles highlighted by Minkler et al. (2008) [36]. Organizational structures within healthcare systems posed challenges for participatory research with providers, notably constrained research time [21, 22]. Roberge-Dao et al. (2019) found that healthcare providers’ current work environment can discourage research involvement, conflicting with researchers’ aspirations amidst organizational barriers [22]. Budget cuts in healthcare impacted human resource management, affecting compensation, benefits, and performance, hindering collaboration with partners for whom research isn’t prioritized [8]. Healthcare partners faced obstacles to research participation due to mission conflicts, reluctance toward random assignment, and rigid payment systems prioritizing productivity over research [49].

##### 2.2. Throughput

###### Lack of guidance and management challenges

This category addresses the lack of guidelines that help manage collaborations with PWDs. Challenges such as conflict of interest, divergent opinions, and incompatible personalities can hinder research efforts when not effectively managed [31]. Various studies showed that lack of comprehensive guidance in participatory research with specific populations exacerbates these challenges [34, 35, 37, 46, 52]. For instance, authors noted a scarcity of models for participatory research involving individuals with intellectual disabilities [46], reproducible details in studies analyzing data from these individuals [37]. Other authors have also noted a predominant emphasis in social sciences on participatory autism research, neglecting other relevant fields [52]. Furthermore, no guidelines have been identified for the collaborative development of clinical care organizations for people with major depressive disorder [35].

###### Challenges in achieving effective involvement of partners

This category covers issues that limit partners’ contributions to research projects. De Wit et al. (2015) noted the challenges researchers encountered in effectively involving partners in research efforts [26]. These challenges included varying expectations regarding participation levels, limited understanding of participatory research and partner roles, and researchers’ struggle to view partners as equal collaborators with shared responsibilities. Additionally, partners may feel uninformed about the time and energy required for research and participatory research development. Moreover, researchers may face conflicts between the data needed for research questions and that generated by partners [42]. Furthermore, participatory research could run the risk of homogeneity when researchers recruit partners similar to themselves, potentially silencing certain voices while amplifying others through participatory methods [42].

##### 2.3. Outcomes

###### Challenges and considerations in participative research

This category highlights the potential drawbacks associated with participatory approach and the involvement of multiple partners. Participatory research was seen as offering numerous benefits but also potentially exacerbating challenges, due to the involvement of multiple partners, as highlighted by various studies [8, 13, 25–28, 31, 34, 39, 40, 42, 43, 46, 51]. These challenges included significant resource commitments, such as additional expenses for partner training, travel, support, and adapting materials to meet partner needs at every stage [8, 13, 31]. Moreover, the participatory approach can increase the workload for researchers, who must provide continuous guidance and support to partners [29, 31]. Furthermore, involving multiple partners can lead to conflicts of interest within the research team, complicating decision-making and project activities [27, 31]. Therefore, it is crucial to carefully assess whether a participatory approach aligns with the specific needs and goals of each research project [30].

## Discussion

The objective of this study was to identify the factors (facilitators and obstacles) that influence the process and results of participatory research in the field of disability studies. As outlined in the results, this mapping review indicates several key factors that contribute to participatory research functioning. This review provides an in-depth insight into how different inputs such as funding opportunities, partners resources and expectations, interact and influence each other within contextual factors to produce different processes and outcomes.

Results from this review identified in literature that key partner resources that are crucial for collaborative project success: researchers and participants training and skills development. Partners’ contributions through expertise, skills, and professional work, are all important inputs. Practicality was emphasized, advocating for structured courses for researchers and participants alike. To the best of our knowledge, there has not yet been a sufficiently exhaustive literature review in the field of disability on the facilitators and barriers to the participatory research process.

Recent literature underscored the benefits of preparing both researchers and participants for collaborative processes [24, 25, 34, 38]. As mentioned in studies, capacity building programs effectively enhanced participatory research through skill development and relationship-building via mentoring or peer support, fostering effective collaboration between community and academic teams [53], as knowledge and skills imbalances among team members as a barrier in participatory research [54]. These studies recommended comprehensive training, creating a conducive environment for participatory research, and ensuring teams were well-equipped with appropriate tools and training before commencing collaborations [53–55].

Including PWDs in research was described as crucial for fostering inclusive projects. Their perspectives and insights were valuable and contributed to the overall success of collaborative projects in disability studies. To ensure meaningful involvement, understanding partners’ specific needs was paramount. Accommodations, specialized training materials, and various communication methods (eg.verbal and non-verbal) were necessary, tailored to different disabilities. Challenges in partnering with PWDs highlighted the importance of creating an inclusive and supportive environment conducive to diverse participation in health research [56]. External constraints like scheduling and limited funding can also pose challenges. Securing adequate funding, including grants supporting partners’ participation, was essential for enabling inclusive research [54, 57]. Our review addresses contextual factors that facilitate partners’ involvement in research, acknowledging the diverse challenges faced by researchers, healthcare providers, and others in collaborative participatory research. These factors significantly influenced partners’ engagement and availability for research collaboration.

Our review provides insight into how different dimensions and processes interact to produce outcomes. The dynamics between different inputs can be affected by contextual factors and managerial variables, as well as their interactions, influencing the effectiveness of performance. In this context, real power sharing, effective communication, mutual learning, and the importance of financial and contextual support were found to be crucial for the success of participatory research in disability studies. This aligns with finding from other studies on the importance of collaboration, trust, and shared decision-making in participatory research [23, 26, 31, 36, 45]. These factors contribute to the overall functioning of participatory research and play a significant role on positive outcomes of participatory research in this field. It is also important to reflect on the limitation of any research project.

### Strengths and weaknesses

This mapping review stands out for its comprehensive coverage of documentation it covered related to participatory research involving PWDs. Thus, significant work was done to synthesize the information present in the various research domains covered. This synthesis of information provides crucial access to knowledge regarding preferred partnership research practices with PWDs, which can contribute to improving research team practices in this area.

However, the study also has some limitations. Due to the numerous terminologies encompassing participatory research, several different terms had to be used in the literature search. Although the concepts of partnership research and participatory research are closely related, partnership research implies greater inclusion of PWDs throughout the research process. In this study, however, the term “participatory research” was preferred due to its greater prevalence in databases. Nevertheless, we discussed articles in which PWDs were involved in the research from beginning to end of the process.

## Conclusion

This study brings together scientific knowledge on facilitators and barriers throughout the partnership research process. This knowledge is essential in a context where researchers are interested in best practices in participatory research with PWDs to contribute to building a more inclusive society. Indeed, an in-depth understanding of the various factors and processes at play in participatory research helps to achieve relevant and positive research results. This study provides valuable insights for future participatory research and research practice in the field of disability studies.

Moving forward, it is essential that future research in this field continues to explore and identify the nuanced dynamics of participatory research processes in order to overcome the various obstacles identified and develop suitable mechanisms for doing so.

## Data Availability

All data produced in the present work are contained in the manuscript

## Acknowledgments

This work was supported by the under grant #295513 of the program “grands défis de société 2021-2013” of the Fonds de recherche du Québec for the Research Team Inclusive Society; under grant #2023-SE7-310780 for the Research Team Support program of the Fonds de recherche du Québec – Société et Culture (FRQSC) (Participation sociales et villes inclusives); under grant #284142 for the doctoral scholarship from FRQSC to MC; under grant Concours 1 for doctoral scholarship from Centre for interdisciplinary research in rehabilitation and social integration (Cirris) to RJ and under grant #296761 for research scholar awards from Fonds de recherche du Québec – Santé (FRQS) to FR.

