## Appendix 1 for "A mapping review of good practices of participatory research for an impactful collaboration in disabilities studies"

**Research Scripts in each database**

**ABI Inform (n=320)**

TI,AB((collaborat* OR partner* OR participatory OR "community based" OR "community centered") NEAR/3 research) OR TI,AB((coproduction* or “co production” or cocreation or “co creation”) NEAR/2 knowledge) OR MESH.exact("Community-Based Participatory Research" OR "cooperative behavior" or “community –institutional relations”) **AND** TI(model or models or guideline* or "guide line*" or guidance or "meta-analy*" OR metaanaly* OR "met analy*" OR metanaly* OR metasynthesis OR "meta synthesis") OR MESH.exact(“guidelines as topic” OR “practice guidelines as topic” or “meta-analysis as topic” OR “meta-analysis” or “models, theoretical” or “models, organizational”)

**Ageline (n=11)**

( ( ((collaborat* OR partner* OR participatory OR "community based" OR "community centered") N3 research) ) OR ( ((coproduction* or “co production” or cocreation or “co creation”) N3 knowledge) ) ) OR ( ("Community-Based Participatory Research" OR "cooperative behavior" or “community –institutional relations”) ) **AND** ( (“guidelines as topic” OR “practice guidelines as topic” or “meta-analysis as topic” OR “meta-analysis” or “models, theoretical” or “models, organizational”) ) OR TI ( (model or models or guideline* or "guide line*" or guidance or "meta-analy*" OR metaanaly* OR "met analy*" OR metanaly* OR metasynthesis OR "meta synthesis") )

**Anthropology plus (n=6)**

( ( (“guidelines as topic” OR “practice guidelines as topic” or “meta-analysis as topic” OR “meta-analysis” or “models, theoretical” or “models, organizational”) ) OR TI ( (model or models or guideline* or "guide line*" or guidance or "meta-analy*" OR metaanaly* OR "met analy*" OR metanaly* OR metasynthesis OR "meta synthesis") ) ) **AND** ( ( ( ((collaborat* OR partner* OR participatory OR "community based" OR "community centered") N3 research) ) OR ( ((coproduction* or “co production” or cocreation or “co creation”) N3 knowledge) ) ) OR ( ("Community-Based Participatory Research" OR "cooperative behavior" or “community –institutional relations”) ) )

**CINAHL Plus with Full Text (n=1095)**

( ( (“guidelines as topic” OR “practice guidelines as topic” or “meta-analysis as topic” OR “meta-analysis” or “models, theoretical” or “models, organizational”) ) OR TI ( (model or models or guideline* or "guide line*" or guidance or "meta-analy*" OR metaanaly* OR "met analy*" OR metanaly* OR metasynthesis OR "meta synthesis") ) ) **AND** ( ( ( ((collaborat* OR partner* OR participatory OR "community based" OR "community centered") N3 research) ) OR ( ((coproduction* or “co production” or cocreation or “co creation”) N3 knowledge) ) ) OR ( ("Community-Based Participatory Research" OR "cooperative behavior" or “community –institutional relations”) ) )

**International bibliography of the social sciences (IBSS) (n=253)**

(TI,AB((collaborat* OR partner* OR participatory OR "community based" OR "community centered") NEAR/3 research) OR TI,AB((coproduction* OR "co production" OR cocreation OR "co creation") NEAR/2 knowledge) OR MESH.exact("Community-Based Participatory Research" OR "cooperative behavior" OR "community --institutional relations")) **AND** (TI(model OR models OR guideline* OR ("guide line" OR "guide lines") OR guidance OR "meta-analy*" OR metaanaly* OR "met analy*" OR metanaly* OR metasynthesis OR "meta synthesis") OR (MESH.exact("guidelines as topic" OR "practice guidelines as topic" OR "meta-analysis as topic" OR "meta-analysis" OR "models, theoretical" OR "models, organizational")))

**Medline (n=196)**

((collaborat* or partner* or participatory or community based or community centered) adj3 research).ti,ab OR ((coproduction* or co production or cocreation or co creation) adj2 knowledge).ti,ab **AND** Community-Based Participatory Research/ or cooperative behavior/ Action research/ or cooperative learning/ **AND** (model or models or guideline* or guide line*or guidance or meta-analy* OR metaanaly* OR met analy* OR metanaly* OR metasynthesis OR meta synthesis).ti OR guidelines as topic/ OR practice guidelines as topic/ or meta-analysis as topic/ OR meta-analysis/ or models, theoretical/ or models, organizational/ meta analysis/ or treatment guideline/

**PsycInfo (n=389)**

((collaborat* or partner* or participatory or community based or community centered) adj3 research).ti,ab OR ((coproduction* or co production or cocreation or co creation) adj2 knowledge).ti,ab **AND** Community-Based Participatory Research/ or cooperative behavior/ Action research/ or cooperative learning/ **AND** (model or models or guideline* or guide line*or guidance or meta-analy* OR metaanaly* OR met analy* OR metanaly* OR metasynthesis OR meta synthesis).ti OR guidelines as topic/ OR practice guidelines as topic/ or meta-analysis as topic/ OR meta-analysis/ or models, theoretical/ or models, organizational/ meta analysis/ or treatment guideline/

**Sociological Abstracts (n=95)**

(TI,AB((collaborat* OR partner* OR participatory OR "community based" OR "community centered") NEAR/3 research) OR TI,AB((coproduction* OR "co production" OR cocreation OR "co creation") NEAR/2 knowledge) OR MESH.exact("Community-Based Participatory Research" OR "cooperative behavior" OR "community --institutional relations")) **AND** (TI(model OR models OR guideline* OR ("guide line" OR "guide lines") OR guidance OR "meta-analy*" OR metaanaly* OR "met analy*" OR metanaly* OR metasynthesis OR "meta synthesis") OR (MESH.exact("guidelines as topic" OR "practice guidelines as topic" OR "meta-analysis as topic" OR "meta-analysis" OR "models, theoretical" OR "models, organizational")))

**Social Science Full Text (n=97)**

( ( (“guidelines as topic” OR “practice guidelines as topic” or “meta-analysis as topic” OR “meta-analysis” or “models, theoretical” or “models, organizational”) ) OR TI ( (model or models or guideline* or "guide line*" or guidance or "meta-analy*" OR metaanaly* OR "met analy*" OR metanaly* OR metasynthesis OR "meta synthesis") ) ) **AND** ( ( ( ((collaborat* OR partner* OR participatory OR "community based" OR "community centered") N3 research) ) OR ( ((coproduction* or “co production” or cocreation or “co creation”) N3 knowledge) ) ) OR ( ("Community-Based Participatory Research" OR "cooperative behavior" or “community –institutional relations”) ) )

**Social Service Abstracts (n=29)**

( ( (“guidelines as topic” OR “practice guidelines as topic” or “meta-analysis as topic” OR “meta-analysis” or “models, theoretical” or “models, organizational”) ) OR TI ( (model or models or guideline* or "guide line*" or guidance or "meta-analy*" OR metaanaly* OR "met analy*" OR metanaly* OR metasynthesis OR "meta synthesis") ) ) **AND** ( ( ( ((collaborat* OR partner* OR participatory OR "community based" OR "community centered") N3 research) ) OR ( ((coproduction* or “co production” or cocreation or “co creation”) N3 knowledge) ) ) OR ( ("Community-Based Participatory Research" OR "cooperative behavior" or “community –institutional relations”) ) )

**Women’s studies International (n=10)**

( ( (“guidelines as topic” OR “practice guidelines as topic” or “meta-analysis as topic” OR “meta-analysis” or “models, theoretical” or “models, organizational”) ) OR TI ( (model or models or guideline* or "guide line*" or guidance or "meta-analy*" OR metaanaly* OR "met analy*" OR metanaly* OR metasynthesis OR "meta synthesis") ) ) **AND** ( ( ( ((collaborat* OR partner* OR participatory OR "community based" OR "community centered") N3 research) ) OR ( ((coproduction* or “co production” or cocreation or “co creation”) N3 knowledge) ) ) OR ( ("Community-Based Participatory Research" OR "cooperative behavior" or “community –institutional relations”) ) )

**Worldwide Political Science Abstracts (n=17)**

(TI,AB((collaborat* OR partner* OR participatory OR "community based" OR "community centered") NEAR/3 research) OR TI,AB((coproduction* OR "co production" OR cocreation OR "co creation") NEAR/2 knowledge) OR MESH.exact("Community-Based Participatory Research" OR "cooperative behavior" OR "community --institutional relations")) **AND** (TI(model OR models OR guideline* OR ("guide line" OR "guide lines") OR guidance OR "meta-analy*" OR metaanaly* OR "met analy*" OR metanaly* OR metasynthesis OR "meta synthesis") OR (MESH.exact("guidelines as topic" OR "practice guidelines as topic" OR "meta-analysis as topic" OR "meta-analysis" OR "models, theoretical" OR "models, organizational")))
