## Appendix 2 for "A mapping review of good practices of participatory research for an impactful collaboration in disabilities studies"

| **Authors** | **Year** | **Areas of disabilities** | **Methodological approaches** | **Partnership approach** |
| --- | --- | --- | --- | --- |
| Abma et al. | 2009 | Intellectual disabilities | Qualitative | Responsive research |
| Baart & Abma, | 2011 | Mental health | Qualitative | Action research / Dialogue Model |
| Bigby & Frawley | 2010 | Intellectual disabilities | Qualitative | Action research / Inclusive research |
| Bigby et al. | 2014 | Intellectual disabilities | Qualitative | Participatory action research (PAR) |
| Camden et al. | 2015 | Multiple | Scoping review | Multiple |
| Clements | 2012 | Mental health | Qualitative | Participatory action research (PAR) / Photovoice |
| Conder et al. | 2011 | Intellectual disabilities | Qualitative | Participatory action research (PAR) |
| de Wit et al. | 2015 | Rheumatism | Mixed methods | First Model |
| Desai et al. | 2019 | Mental health | Qualitative | Participatory research |
| Ehde et al. | 2013 | Physical disabilities | Literature review / Mixed methods | Participatory Action Research (PAR) / Community-based participatory research (CBPR) |
| Fletcher-Watson et al. | 2019 | Autism | Qualitative^[1]^ | Participatory research |
| Frounfelker et al. | 2012 | Mental health | Qualitative | Academic-provider research partnerships |
| Garcia‐Iriarte et al. | 2009 | Intellectual disabilities | Mixed methods | Participatory action research (PAR) |
| Hassouneh et al. | 2011 | Physical disabilities (mobility or sensory impairment | Mixed methods | Community-based participatory research (CBPR) |
| Heffron et al. | 2018 | Intellectual and developmental disabilities. | Qualitative | Participatory action research (PAR) / Photovoice |
| Jinks et al. | 2009 | Knee pain | Qualitative | INVOLVE (model for involvement in research) |
| Jivraj et al. | 2014 | Autism spectrum disorder and neurodevelopmental disorders | Scoping review | Participatory research (PR) |
| Khodyakov et al. | 2011 | Mental Health | Mixed methods | Conceptual model based on Community-based participatory research (CBPR) and the partnership synergy model ((Lasker et al. 2001; Weiss et al. 2002) |
| King et al. | 2008 | Physical disabilities, developmental disabilities, and/or communication needs^[2]^ | Qualitative | Four research program operating models :   - “Clinician- Researcher” Skill Development Model - Clinician and Researcher Program Evaluation Model / Researcher- Led - Knowledge Generation Model - Knowledge Conduit Model |
| Kramer et al. | 2011 | Intellectual disabilities | Mixed methods | Participatory action research (PAR) |
| Lannin et al. | 2021 | Brain injury | Qualitative | Citizen Jury method of participatory research. |
| Lincoln et al. | 2015 | Mental Health | Qualitative | Community-Based Participatory Research (CBPR) |
| Littlechild et al. | 2015 | Dementia | Qualitative | Co-research approach |
| MacLeod | 2019 | Autism | Systematic Review | Participatory research |
| McDonald & Stack | 2016 | Developmental disabilities | Qualitative | Community-based participatory research (CBPR) |
| Minkler et al. | 2008 | Disabilities at large | Mixed methods | Community-based participatory research (CBPR) |
| Morton | 2012 | Non-visible handicaps, specifically those with epilepsy | Qualitative | Participatory action research (PAR) / Photovoice |
| Moule & Davies | 2016 | Mental Health | Qualitative | Not specified |
| Nicolaidis et al. | 2019 | Autism | Qualitative | Community-based participatory research (CBPR) / Consultative model (community advisory board) |
| Nicolaidis et al. | 2011 | Autism | Qualitative | Community-based participatory research (CBPR) |
| Nierse & Abma, | 2011 | Intellectual disabilities | Qualitative | Responsive research |
| Ottmann et al. | 2009 | Disabilities at large | Qualitative | Participatory Action Research (PAR) |
| Ottmann et al. | 2011 | Older people with complex health issues | Qualitative | Coproduction Approach /  Participatory Action Research / Co-operative Inquiry (CI) |
| Povee et al. | 2014 | Intellectual disabilities | Qualitative | Participatory research approaches / Photovoice |
| Riemer et al. | 2012 | Mental Health | Qualitative | University-Practice Partnership (community-based participatory partnerships) |
| Roberge-Dao et al. | 2019 | Rehabilitation | Mixed methods | Integrated knowledge translation (IKT) |
| Schneider | 2012 | Mental Health | Qualitative | Participatory action research (PAR) |
| Stevenson | 2014 | Down Syndrome | Qualitative | Participatory action research |
| Stolee et al. | 2011 | Dementia | Mixed methods | Consensus Workshop |
| Tanner | 2012 | Dementia | Qualitative | Not specified |
| Tavecchio et al. | 2019 | intellectual disabilities | Mixed methods | Participatory Peer Research (PPR) |
| Thoft et al. | 2020 | Dementia | Qualitative | Partners in Projects / Authentic partnerships / Balanced Participation Model / |

1. Present outcomes from a series of UK seminar series
2. children and young people
